# Genetics of Congenitally Corrected Transposition of the Great Arteries: next generation sequencing shows a mutation load effect for 156 genes involved in cardiac patterning

**DOI:** 10.1101/2023.09.11.23293772

**Authors:** Yasmine Benadjaoud, Sabina Benko, Fabienne Jabot Hanin, Enrique Audain, Jean François Deleuze, Anne Boland, Christine Bole, Marc Bras, Fanny Bajolle, Morgan Derouk, Christopher T. Gordon, Marc Phillip Hitz, Damien Bonnet, Stanislas Lyonnet

## Abstract

Congenital heart disease (CHD) is a major public health issue. It is considered as a major cause of infant mortality today.

We focused on a complex and rare CHD, the congenitally corrected transposition of the great arteries (CCTGA).

The aim of this study was to identify the genes that control the alignment of cardiac chambers that are abnormal in CCTGA to further understand the mechanism of this CHD.

We analyzed a cohort of 43 CCTGA cases (41 sporadic cases and 2 familial cases) of isolated CCTGA by next generation sequencing analysis (whole-genome sequencing and whole-exome sequencing).

Under the hypothesis of Mendelian model including *de novo* genomic alterations, no major gene effect involved in the disease could be identified.

Under the hypothesis of a complex model of inheritance, we highlighted a group of 156 cardiac mutated genes in our patients, in which we found a significant enrichment of rare variants in patients compared to controls in a replication cohort. The highly heterogeneous combinations of susceptibility rare variants, mostly inherited from the healthy mother and father respectively, are correlated with the CCTGA phenotype, any given deleterious variant combination within the CCTGA gene set being specific to the affected individual.

Taken together, the cases could be explained by a mutation load of segregated alleles at loci mostly involved in heart tube *looping*, outflow tract morphogenesis and establishment of left-right asymmetry. Our data suggest that CHD are not Mendelian traits, but rather of polygenic origin. In the majority of cases, both parents harbour a set of susceptibility alleles for the CHD that are inherited by the offspring in a combination that confers the risk of CCTGA.

## Introduction

Congenital heart disease (CHD) is the most common human birth defects, occurring in 8/1000 live births ^1 2^. It is still considered as a major cause of infant mortality today^3^.

The defects comprise an assortment of structural malformations, ranging from minor septal defects to complex life-threatening malformations, which require highly specialized medical care.

Some category of CHDs is due to a misalignment of cardiac chambers (ventricles and great arteries).

We focus on a complex and rare CHD: the congenitally corrected transposition of the great arteries (CCTGA). Its incidence is 1/33000 live birth, accounting for 0.05% of all CHDs ^4^. It implies both atrio-ventricular discordance (the right atrium is emptying into the left ventricle (LV), and left atrium emptying into the right ventricle (RV)) and ventriculo-arterial discordance (the anterior and leftward positioned aorta arises from the RV, while the pulmonary artery, positioned posteriorly and rightward, arises from the LV) ^5^ .

Embryologically, CCTGA is considered to be consequential to the abnormal leftward looping of the primitive heart tube, which results in the morphological LV being positioned to the right of the morphological RV ^6^.

The genetic bases of the CCTGA are still unknown ^7^.

It is most often of sporadic appearance. However, there are some rare familial cases reported in the literature ^7 8^. These reported pedigrees suggest a common pathogenetic pathway involving laterality genes in the pathophysiology of CCTGA. Globally, the risk for a recurrence in subsequent pregnancy for healthy parents with a first child with a CHD is estimated at 1-3% ^9^.

The aim of this study is to better understand the genetic mechanisms that control the alignment of cardiac chambers and that lead to the CCTGA.

We performed whole-genome and whole-exome sequencing in a cohort of CCTGA. Our data favour a polygenic origin of the disease in the majority of cases, where there is addition of pathogenic variants inherited from the father and the mother, and this addition of variants lead to CCTGA.

## Material and methods

### 1. The CARREG Collection

All the DNAs of the patients and their parents were obtained thanks to a bio-collection of congenital heart defects in children, based on the collection of DNA by blood sampling. This DNA library, called CARREG (CARdiac congenital defects and REGulation genes) was created by Pr. Damien Bonnet from the M3C-Necker pediatric cardiology department of Necker-Enfants Malades Hospital, and contains more than 4000 referenced samples of patients with different congenital heart defects.

### 2. DNA extraction

Patients and parents DNAs were extracted from a leukocyte pellet. It is an extraction method without organic solvent, by sodium dodecyl sulphate (SDS) deionization, proteinase K digestion, denaturation and protein precipitation by saturated NaCl. Precipitation of the DNA is carried out by adding absolute ethanol at -20 ° C. Excess NaCl is removed by addition of 70% ethanol at 4° C. Finally, the DNA is suspended in the water.

### 3. Next Generation Sequencing (NGS)

#### a) Whole Genome Sequencing: WGS

The DNAs were purified on AMICON® Ultra columns and their quality verified on an agarose gel. Libraries were created after ultrasonic DNA fragmentation (Covaris E210) using the Illumina TruSeq v3 kit. Complete genome sequencing (WGS) was performed in paired end (2×100 bases) on an Illumina HiSeq2000 platform. The mean depth of sequencing was on the order of 30 reads, except for coverage risks due to the intrinsic quality of the sample.

#### b) Whole Exome Sequencing: WES

8 µg of genomic DNA has been purified on AMICON® Ultra columns. Its quality was verified on a 2% agarose gel and the concentration of each sample was measured by EXPOSE®. For the production of the exome library, 3 μg of genomic DNA was used with the Agilent SureSelect v5 all Human Exon Kits. The exon multiplex bank was sequenced on a HISeq2500 machine (Illumina) in “paired-end 76 + 76” mode.

### 4. Polymerase Chain Reaction (PCR)

PCR (Polymerase Chain Reaction) reactions, optimized on control DNA, were performed using GoTaq DNA polymerase (Promega®). The final volume of PCR mixture was 25 µl. PCR reactions were performed using the DNA of the index individuals extracted from blood or saliva, in an Eppendorf® thermal cycler. PCR products were visualized on a 1% agarose gel with bromophenol blue as a loading buffer. To verify the size of the amplicons we used the Roche Applied Science® DNA VIII Molecular Weight Marker.

### 5. Sanger sequencing

The amplicons obtained by PCR were subjected to sequencing by the Sanger method. For this, the PCR products were first purified of excess primers and nucleotides using ExoSap-IT (Affimetrix®): 5µl of PCR product+ 2µl of ExoSap. Sanger sequencing makes it possible to read the DNA sequence using fluorophores. When a ddNTP is added to the DNA chain, it stops its elongation and emits a coloured light that is specific to it: we can then identify the nucleotide. These multiple elongations are performed simultaneously on many fragments, thus obtaining the complete sequence. Thus, we used the Big Dye® Terminator v3.1 Cycle Sequencing kit (Applied Biosystems®) to generate the fluorescent fragments, which are then purified on Sephadex G-50 Fine gel to remove low molecular weight impurities. The product is then deposited on the Applied Biosystems 3500xl Genetic Analyser automatic sequencer. The results are analyzed using Sequencing Analysis software (Sequencing Analysis V6.0). The sequences obtained are finally compared with the reference sequence of the human genome (hg19 assembly).

### 6. Cohort description

#### Primary cohort

Patients with CCTGA were recruited in Necker-Enfants Malades Hospital thanks to the CARREG program. A total of 18 trios (2 parents without CCTGA and 1 child), 17 families with 2 parents, sibs and 1 child affected, 4 duos (1 parent without CCTGA and 1 child), 1 family with a parent and a child both affected, 1 family with 2 affected were recruited.

The inclusion criteria were isolated CCTGA patients without extra-cardiac anomalies. Associated cardiac anomalies were accepted.

502 ethnically matched internal “controls” were also included in the study. They were patients studied at the Imagine Institute, Paris, France, and suffering mainly from immune related diseases.

#### Replication cohort

We used a cohort from 86 unrelated CCTGA patients and 5163 controls from the Wellcome Sanger Institute for replication, all from Caucasian origin. The individuals were sequenced with Agilent SureSelect capture kit V3, V4 or V5. Cases and controls were aligned with BWA software and jointly called with GATK haplotypecaller standard pipeline. Variant effect predictor tool (VEP, API 94) was used to annotate the dataset, and all quality controls and analyses were performed in HAIL 0.2.

Only bi-allelic and exonic variants were considered, with a mean DP > 15, a mean GQ > 30 and call_rate > 0.95 over the whole cohort. Individual genotypes were set up to missing if Allelic Balance < 20% or > 80% for heterozygous.

### 7. Bioinformatics analysis

The downstream analyses of primary cohort’s sequencing data were performed at the Bioinformatics platform of the University of Paris, France. After demultiplexing, paired-end sequences were aligned to the reference human genome (GRCh37) with the Burrows-Wheeler Aligner ^10^. The mean depth of coverage obtained for each sample was greater than 150× for WES and greater than 30x for WGS, and more than 96% of the exome was covered at least 15× in each WES or WGS.

Downstream processing was carried out with the Genome Analysis Toolkit (GATK) ^11^, SAM tools ^12^, and Picard according to documented best practices. GATK Haplotype caller was used for variant calling.

Patients-based *de novo* variants analysis was performed with an in-house software system (PolyWeb) which allows annotation and filtering of variants according to relevant genetic models. All *de novo* variants were then confirmed by Sanger sequencing.

Cases and controls of primary cohort were jointly called with GATK Haplotype caller. We used Variant Quality Score Recalibration (VQSR) to filter the joint VCF file following GATK recommendations for parameter settings: HapMap 3.3, Omni 2.5, dbSNP 138, 1000 Genome phase I for SNPs training sets, and Mills- and 1000 Genome phase I data for indels, and we kept only variants with a VQSLOD score above the tranche sensitivity threshold of 99%. Individual genotypes with a DP <8x, GQ <20 and minor allele ratio < 20% were set to missing.

### 8. Ethnicity inference

For selection of individuals from European or North African ancestry, we extracted 60 000 single nucleotide polymorphisms with a MAF > 5% in 1000 Genomes Project Phase3 covered by the WES capture kit and performed a Principal Component Analysis on 1000 Genomes data with projection of patients and controls on the graph, thanks to Plink1.9 software ^13 14^.

43 patients from the primary cohort and 502 controls were first selected based on this analysis to include a maximum number of patients **(supplementary figure 1).** The individual number of synonymous variants (assumed not to be associated with the phenotype) was compared between cases and controls to check the comparability of data **(supplementary figure 2).**

### 9. Association analysis

After quality filtering, all variants from the primary cohort were annotated using SnpEff ^15^. Only variants with a global call rate > 95% and annotated as missense, frameshift, disruptive inframe deletion/insertion, splice donor or acceptor, or stop gain as most severe consequence among analysed transcripts, and with a minor allele frequency below 0.1% in gnomAD database were kept in the analysis. Missense variants pathogenicity were analysed by an *in silico* pathogenicity criteria based on the CADD score ^16^ and/or on SIFT and PolyPhen softwares 17.

The evolutionary conservation at each residue was also investigated using multiple sequence alignments from the UCSC genome browser.

### 10. Family-based design

We investigated potential risk alleles transmission in the 43 families through the Rare-Variant Generalized Disequilibrium Test ^18^. This method is an extension of the Transmission Disequilibrium Test (TDT) allowing the grouping of information across multiple rare variants distributed within a genomic region. In this analysis, we considered rare functional variants (as previously defined) with or without a CADD score >15, and aggregated them at the gene level.

### 11. Case - control Design

An enrichment in rare variants was investigated at the individual gene level and at cardiac genesets level. Two different methods were used: the CAST method comparing the number of carriers of selected variants between cases and controls to test if a gene or group of genes is preferentially mutated among cases, and a logistic regression method, comparing the number of alleles in the considered genes or genesets between the 2 groups of individuals, to test if an accumulation of variants in a gene or group of genes could explain the phenotype.

### 12. Cardiac genes

We selected a list of cardiac genes, described in the literature to play a role in cardiac morphogenesis in human and mouse. We used Gene Ontology (GO) ^19 20 21^ Consortium and Mouse Genome Informatics (MGI) ^22 23^. The terms used in GO were: heart morphogenesis, heart tube formation, outflow tract development, directing laterality specification. Genes expressed during embryonic heart development were also added. A total of 1708 genes were selected.

## Results

We sequenced by NGS a cohort of 43 families with 44 patients with CCTGA, from the CARREG program.

The cohort was composed of: 17 families with 2 healthy parents and siblings, 18 trios (healthy parents-patient), 4 duos (healthy mother-patient), 2 simplex cases (single patient) and 2 family cases: 1 case of recurrence of CCTGA in siblings (affected brother and sister) and 1 case of vertical transmission of CCTGA (affected father and patient).

The cohort included 18 girls and 26 boys (the boy/girl sex ratio is 1.5).

All patients had isolated CCTGA, with no extracardiac anomalies. The CCTGA could be simple or associated with other cardiac anomalies such as ventricular septal defects, Ebstein morphology of the tricuspid valve, or pulmonary stenosis.

The demographic and anatomical description of the cohort is shown in **Table (1)**.

| N° | Family code | Sex patient | Complete cardiologic diagnosis | Cardiac arrhythmias |
| --- | --- | --- | --- | --- |
| 1 | A001 | F | SS, mesocardia, CCTGA, large VSD, pulmonary atresia | 0 |
| 2 | A002 | M | SS, levocardia, CCTGA, Ebstein disease | WPW |
| 3 | A003 | M | SS, levocardia, CCTGA, Ebstein disease | CAVB |
| 4 | A004 | M | SS, levocardia, CCTGA | 0 |
| 5 | A005 | F | SS, dextrocardia, CCTGA, VSD, pulmonary atresia | postoperative CAVB |
| 6 | A006 | M | SS, mesocardia, CCTGA, VSD, pulmonary atresia | 0 |
| 7 | A007 | F | SS, levocardia, CCTGA, Ebstein disease | 0 |
| 8 | A008 | F | SS, levocardia, CCTGA, VSD, mitral cleft, Ebstein disease | CAVB |
| 9 | A009 | M | SS, levocardia, CCTGA, VSD, pulmonary stenosis | 0 |
| 10 | A010 | F | SS, mesocardia, CCTGA, large VSD, pulmonary stenosis, bicuspid aortic valve | WPW |
| 11 | A011 | M | SS, levocardia, CCTGA, VSD | 0 |
| 12 | A012 | F | SS, levocardia, CCTGA, multiple VSDs, coarctation of the aorta | 0 |
| 13 | A013 | M | SS, levocardia, CCTGA, multiple VSDs, coarctation of the aorta, tricuspid straddling | 0 |
| 14 | A014 | M | SS, levocardia, CCTGA | CAVB |
| 15 | A015 | M | SS, levocardia, CCTGA | 0 |
| 16 | A016 | M | SS, levocardia, CCTGA | 0 |
| 17 | A017 | M | SS, levocardia, CCTGA | 0 |
| 18 | A018 | M | SS, levocardia, CCTGA, restrictive VSD, pulmonary stenosis | CAVB |
| 19 | A019 | F | SS, mesocardia, CCTGA, VSD, pulmonary stenosis | 0 |
| 20 | A020 | M | SS, levocardia, CCTGA, VSD, hypoplasia of RV | JT, CAVB |
| 21 | A021 | M | SS, levocardia, CCTGA, hypoplasia of RV, VSD, pulmonary stenosis | CAVB |
| 22 | A022 | M | SS, levocardia, CCTGA, pulmonary stenosis | 0 |
| 23 | A023 | F | SS, levocardia, CCTGA, VSD | CAVB |
| 24 | A024 | F | SS, levocardia, CCTGA, restrictive VSD, Ebstein disease | 0 |
| 25 | A025 | F | SS, dextrocardia, CCTGA, VSD, pulmonary stenosis | 0 |
| 26 | A026 | M | SS, levocardia, CCTGA, large multiple VSDs | 0 |
| 27 | A027 | F | SS, levocardia, CCTGA | WPW |
| 28 | A028 | M | SI, dextrocardia, CCTGA, VSD, pulmonary stenosis, mitral straddling | 0 |
| 29 | A029 | M | SS, mesocardia, CCTGA, VSD, pulmonary atresia | 0 |
| 30 | A030 | M | SS, mesocardia, CCTGA, large VSD, pulmonary atresia | 0 |
| 31 | A031 | F | SS, levocardia, CCTGA | CAVB |
| 32 | A032 | F | SS, levocardia, CCTGA | 0 |
| 33 | A033 | F | SS, levocardia, CCTGA, VSD | CAVB |
| 34 | A034 | M | SS, dextrocardia, CCTGA | 0 |
| 35 | A035 | F | SS, dextrocardia, CCTGA, VSD | 0 |
| 36 | A036 | M | SS, levocardia, CCTGA, hypoplasia of RV, 2 large VSDs, LSVC | 0 |
| 37 | A037 | F | SS, dextrocardia, CCTGA, VSD, coarctation of the aorta | 0 |
| 38 | A038 | M | SS, levocardia, CCTGA, VSD, tricuspid straddling | 0 |
| 39 | A039 | F | SS, levocardia, CCTGA | CAVB |
| 40 | A040 | M | SI, dextrocardia, CCTGA, hypoplasia of LV | 0 |
|  |  | F | SS, levocardia, CCTGA | 0 |
| 41 | A041 | M | SS, dextrocardia, CCTGA | 0 |
| 42 | A042 | M | SS, levocardia, CCTGA | 0 |
| 43 | A043 | M | SS, levocardia, CCTGA, VSD | 0 |

CAVB: complete atrio-ventricular block, CCTGA: congenitally corrected transposition of the great arteries, JT: junctional tachycardia, LSVC: left superior vena cava, LV: left ventricle, RV: right ventricle, SS : situs solitus, SI: situs inversus , VSD : ventricular septal defect, WPW: Wolf Parkinson White

### *de novo* analysis

Applying the *de novo* mode of inheritance in Polyweb, we selected a list of 20 genes with a *de novo* variant (loss of function or missense) in 17 different families that meet the selection criteria for frequency and nature of variation. 6 variants are loss of function (frameshift insertion or deletion, splice acceptor donor, start-stop lost, STOP gain) and 14 are missense variants.

This list of genes with the description of the variant, its CADD score and the probability to loss of function intolerance (PLI score) is presented in **table (2)**. The presence of another variant inherited from one of the parents in each gene is also detailed.

**Table 2:**
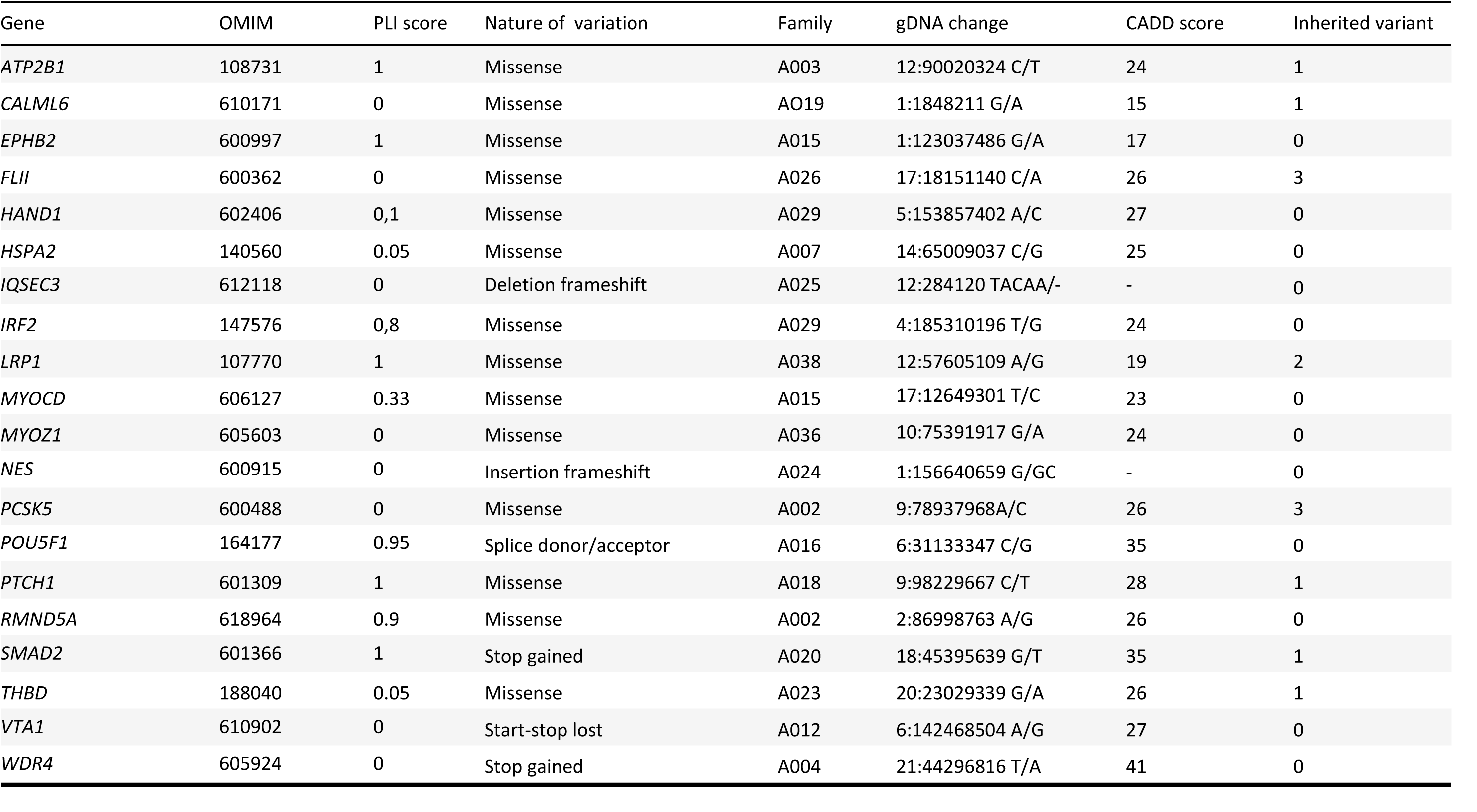
*de novo* variants in CCTGA cohort.

| Gene | OMIM | PLI score | Nature of variation | Family | gDNA change | CADD score | Inherited variant |
| --- | --- | --- | --- | --- | --- | --- | --- |
| *ATP2B1* | 108731 | 1 | Missense | A003 | 12:90020324 C/T | 24 | 1 |
| *CALML6* | 610171 | 0 | Missense | A019 | 1:1848211 G/A | 15 | 1 |
| *EPHB2* | 600997 | 1 | Missense | A015 | 1:123037486 G/A | 17 | 0 |
| *FLII* | 600362 | 0 | Missense | A026 | 17:18151140 C/A | 26 | 3 |
| *HAND1* | 602406 | 0,1 | Missense | A029 | 5:153857402 A/C | 27 | 0 |
| *HSPA2* | 140560 | 0.05 | Missense | A007 | 14:65009037 C/G | 25 | 0 |
| *IQSEC3* | 612118 | 0 | Deletion frameshift | A025 | 12:284120 TACAA/- | - | 0 |
| *IRF2* | 147576 | 0,8 | Missense | A029 | 4:185310196 T/G | 24 | 0 |
| *LRP1* | 107770 | 1 | Missense | A038 | 12:57605109 A/G | 19 | 2 |
| *MYOCD* | 606127 | 0.33 | Missense | A015 | 17:12649301 T/C | 23 | 0 |
| *MYOZ1* | 605603 | 0 | Missense | A036 | 10:75391917 G/A | 24 | 0 |
| *NES* | 600915 | 0 | Insertion frameshift | A024 | 1:156640659 G/GC | - | 0 |
| *PCSK5* | 600488 | 0 | Missense | A002 | 9:78937968A/C | 26 | 3 |
| *POU5F1* | 164177 | 0.95 | Splice donor/acceptor | A016 | 6:31133347 C/G | 35 | 0 |
| *PTCH1* | 601309 | 1 | Missense | A018 | 9:98229667 C/T | 28 | 1 |
| *RMND5A* | 618964 | 0.9 | Missense | A002 | 2:86998763 A/G | 26 | 0 |
| *SMAD2* | 601366 | 1 | Stop gained | A020 | 18:45395639 G/T | 35 | 1 |
| *THBD* | 188040 | 0.05 | Missense | A023 | 20:23029339 G/A | 26 | 1 |
| *VTA1* | 610902 | 0 | Start-stop lost | A012 | 6:142468504 A/G | 27 | 0 |
| *WDR4* | 605924 | 0 | Stop gained | A004 | 21:44296816 T/A | 41 | 0 |

Among the 20 genes with *de novo* variation, 15 have a cardiac function described in the literature (GO, MGI). These genes are: *ATP2B1, EPHB2, FLII, HAND1, HSPA2, LRP1, MYOCD, PCSK5, POU5F1, PTCH1, RMND5A, SMAD2, THBD, VTA1 and WDR4*.

If we consider that 1708 genes / 19370 protein-coding whole genome genes have a role cardiac development of function, and in our cohort 15 genes / 20 genes with *de novo* variant have also cardiac function, by performing a hypergeometric test, we found a significant enrichment of cardiac genes in the *de novo* genes (p<10E-10).

To investigate possible variant enrichment in *de novo* genes, we analyzed variants annotated by SnpEff as disruptive inframe insertion/deletion, frameshift, missense, splice acceptor, splice donor, stop gained, stop lost.

We analyzed the 26 families in whom no *de novo* variant was found. We do not find any enrichment of variants in the 27 cases compared to the 502 controls with a p value=0.83 for a MAF <1%. The same results are seen on the list of 15 genes with a cardiac function.

Considering the enrichment in cardiac genes in *de novo* analysis, we focused on the list of 1708 cardiac genes.

We thus made a comparative analysis of the number of variants in these 1708 genes in patients and controls.

We observed a trend towards an enrichment in the number of very rare variants (MAF <0.01%) in cardiac genes in patients compared to controls (p=0.025). By adjusting for the first 5 principal components (PC), the significance of this enrichment increases (p value 0.005).

These results are shown in **Table (3)**.

**Table 3:** statistical analysis case/control of variants in the 1708 cardiac genes in CCTGA cohort.

|  | Number of variants | Ncarriers_controls / 502 | Ncarriers_cases/ 43 | Total alleles Nb controls | Total alleles Nb cases | Alleles Nb / control | Alleles Nb /case | pvalueLRT | pvalueWald |
| --- | --- | --- | --- | --- | --- | --- | --- | --- | --- |
| **MAF1%, CADD>15** | 13853 | 502 | 43 | 24016 | 2152 | 47.8 | 50 | 0.44 | 0.43 |
| **MAF 0.1%, CADD>15** | 9951 | 502 | 43 | 11586 | 1076 | 23.1 | 25 | 0.21 | 0.21 |
| **MAF0.01%, CADD >15** | 6326 | 500 | 43 | 6252 | 627 | 12.5 | 14.6 | **0.025** | **0.022** |
| **MAF<0.01%, CADD>15**<br>**Adjusted on the 5th first PCs** |  |  |  |  |  |  |  | **0.005** | **0.004** |

To go further, we looked among the 1708 cardiac genes, if some played a more important role than the others and presented a higher number of variants in the cases than in the controls. To do this, we performed statistical analysis using the CAST method and logistic regression tests.

We analyzed 2 types of variants: PTV (protein truncating variant) represented frameshift_variant, stop_gained, splice_donor_variant, splice_acceptor_variant, and PAV (protein altering variant) represented missense variant, inframe deletion, inframe insertion, start lost, stop retained variant, stop lost, protein altering variant, start retained variant .

No gene passed the significance threshold.

### Analysis by group of genes: the polygenic model

By analyzing the families of patients, among the 1708 cardiac genes, a group of 156 genes seemed to group interesting variants meeting the defined selection criteria.

This list of 156 genes is shown in **Table (4)**.

**Table 4:**
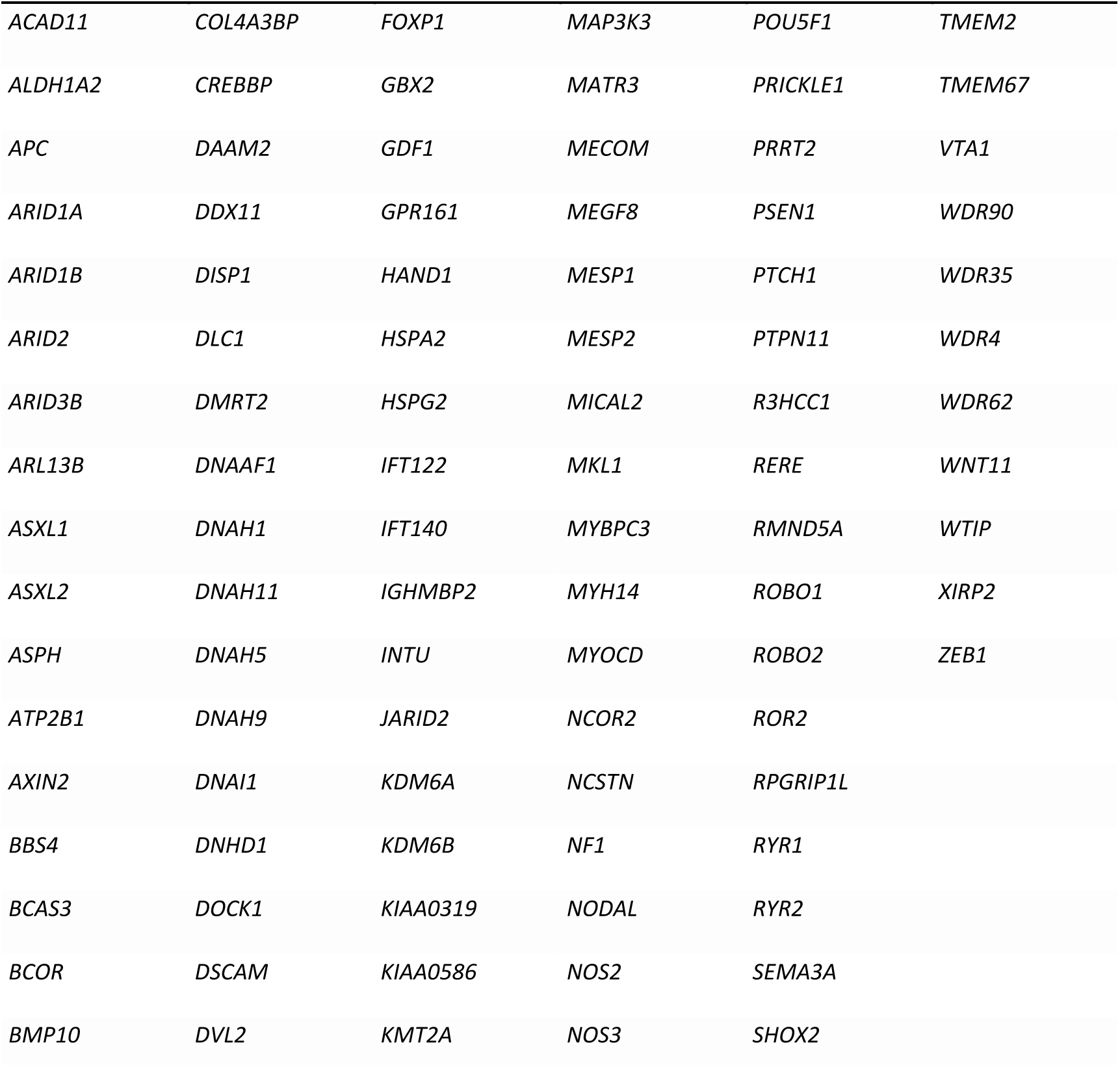
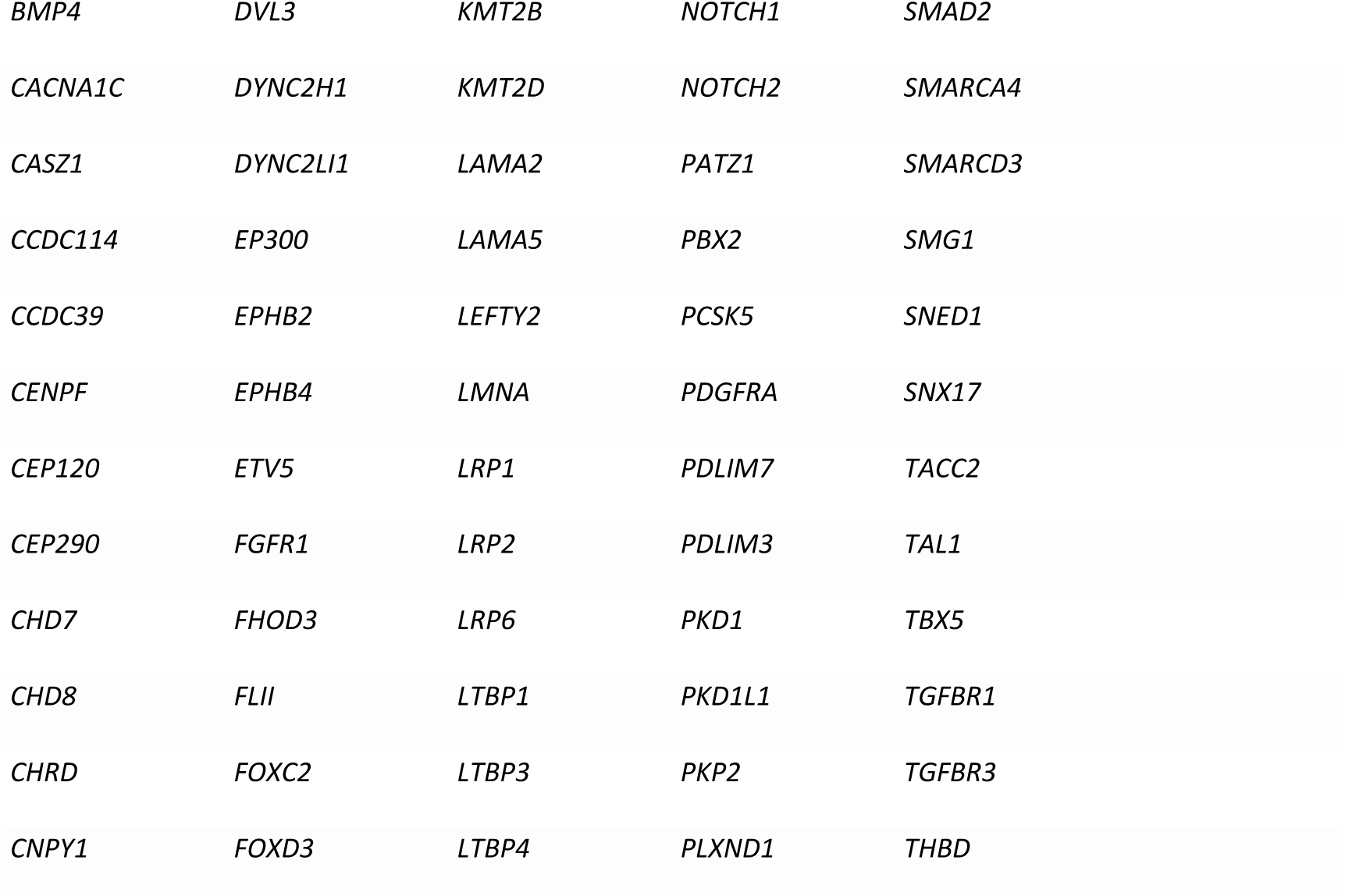
List of 156 cardiac genes with pathogenic variant in CCTGA cohort.

|  |  |  |  |  |  |
| --- | --- | --- | --- | --- | --- |
| *ACAD11* | *COL4A3BP* | *FOXP1* | *MAP3K3* | *POU5F1* | *TMEM2* |
| *ALDH1A2* | *CREBBP* | *GBX2* | *MATR3* | *PRICKLE1* | *TMEM67* |
| *APC* | *DAAM2* | *GDF1* | *MECOM* | *PRRT2* | *VTA1* |
| *ARID1A* | *DDX11* | *GPR161* | *MEGF8* | *PSEN1* | *WDR90* |
| *ARID1B* | *DISP1* | *HAND1* | *MESP1* | *PTCH1* | *WDR35* |
| *ARID2* | *DLC1* | *HSPA2* | *MESP2* | *PTPN11* | *WDR4* |
| *ARID3B* | *DMRT2* | *HSPG2* | *MICAL2* | *R3HCC1* | *WDR62* |
| *ARL13B* | *DNAAF1* | *IFT122* | *MKL1* | *RERE* | *WNT11* |
| *ASXL1* | *DNAH1* | *IFT140* | *MYBPC3* | *RMND5A* | *WTIP* |
| *ASXL2* | *DNAH11* | *IGHMBP2* | *MYH14* | *ROBO1* | *XIRP2* |
| *ASPH* | *DNAH5* | *INTU* | *MYOCD* | *ROBO2* | *ZEB1* |
| *ATP2B1* | *DNAH9* | *JARID2* | *NCOR2* | *ROR2* |  |
| *AXIN2* | *DNAI1* | *KDM6A* | *NCSTN* | *RPGRIP1L* |  |
| *BBS4* | *DNHD1* | *KDM6B* | *NF1* | *RYR1* |  |
| *BCAS3* | *DOCK1* | *KIAA0319* | *NODAL* | *RYR2* |  |
| *BCOR* | *DSCAM* | *KIAA0586* | *NOS2* | *SEMA3A* |  |
| *BMP10* | *DVL2* | *KMT2A* | *NOS3* | *SHOX2* |  |

|  |  |  |  |  |
| --- | --- | --- | --- | --- |
| *BMP4* | *DVL3* | *KMT2B* | *NOTCH1* | *SMAD2* |
| *CACNA1C* | *DYNC2H1* | *KMT2D* | *NOTCH2* | *SMARCA4* |
| *CASZ1* | *DYNC2LI1* | *LAMA2* | *PATZ1* | *SMARCD3* |
| *CCDC114* | *EP300* | *LAMA5* | *PBX2* | *SMG1* |
| *CCDC39* | *EPHB2* | *LEFTY2* | *PCSK5* | *SNED1* |
| *CENPF* | *EPHB4* | *LMNA* | *PDGFRA* | *SNX17* |
| *CEP120* | *ETV5* | *LRP1* | *PDLIM7* | *TACC2* |
| *CEP290* | *FGFR1* | *LRP2* | *PDLIM3* | *TAL1* |
| *CHD7* | *FHOD3* | *LRP6* | *PKD1* | *TBX5* |
| *CHD8* | *FLII* | *LTBP1* | *PKD1L1* | *TGFBR1* |
| *CHRD* | *FOXC2* | *LTBP3* | *PKP2* | *TGFBR3* |
| *CNPY1* | *FOXD3* | *LTBP4* | *PLXND1* | *THBD* |

Each patient carries an association of variants (between 3 and 10 variants), inherited from the father and the mother, in addition of a *de novo* variant.

The genotype of each of the 43 families in the CCTGA cohort (inherited and *de novo* variants) is represented by the pedigrees in **Figure (1)**.

**Figure (1):**
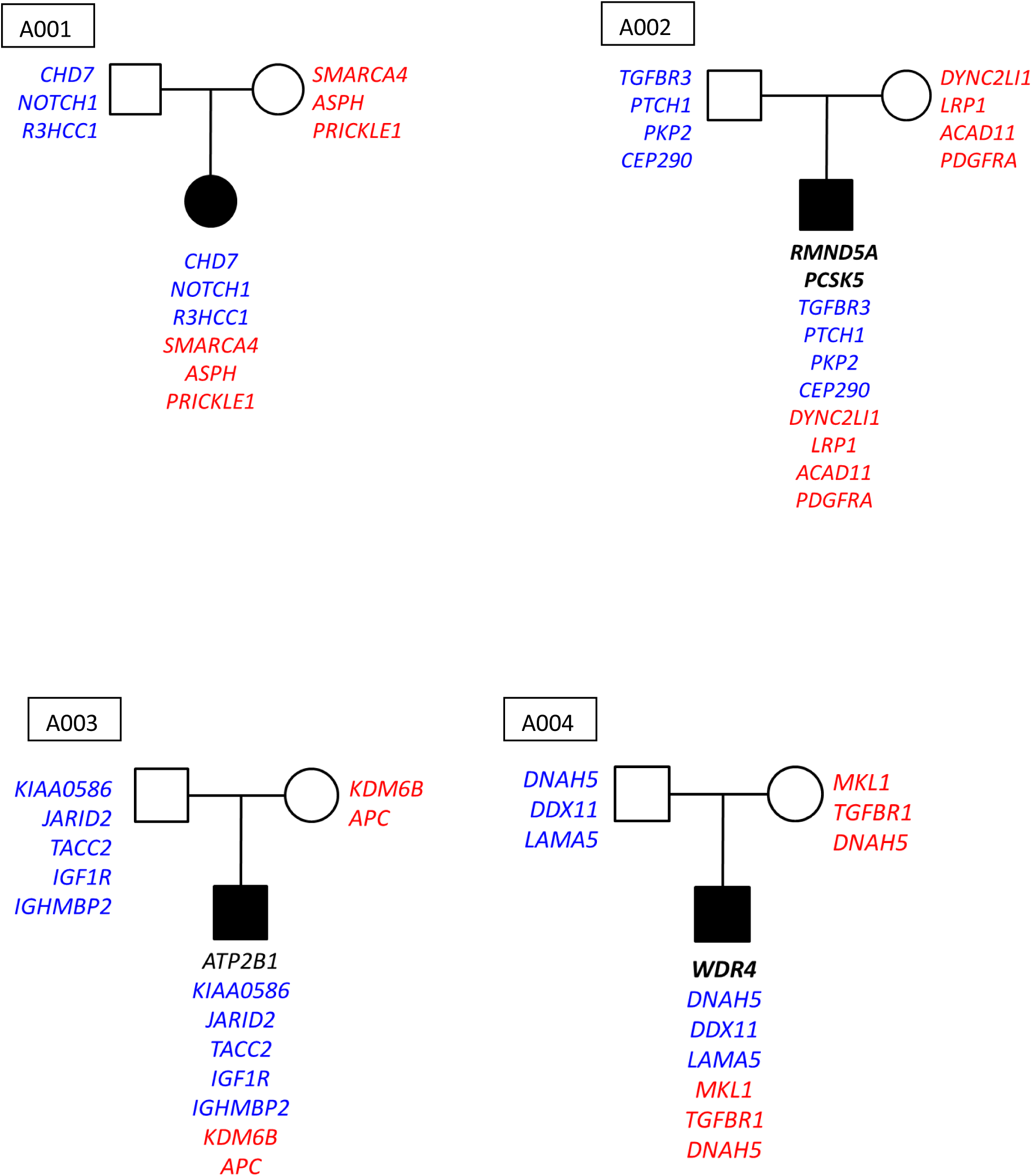
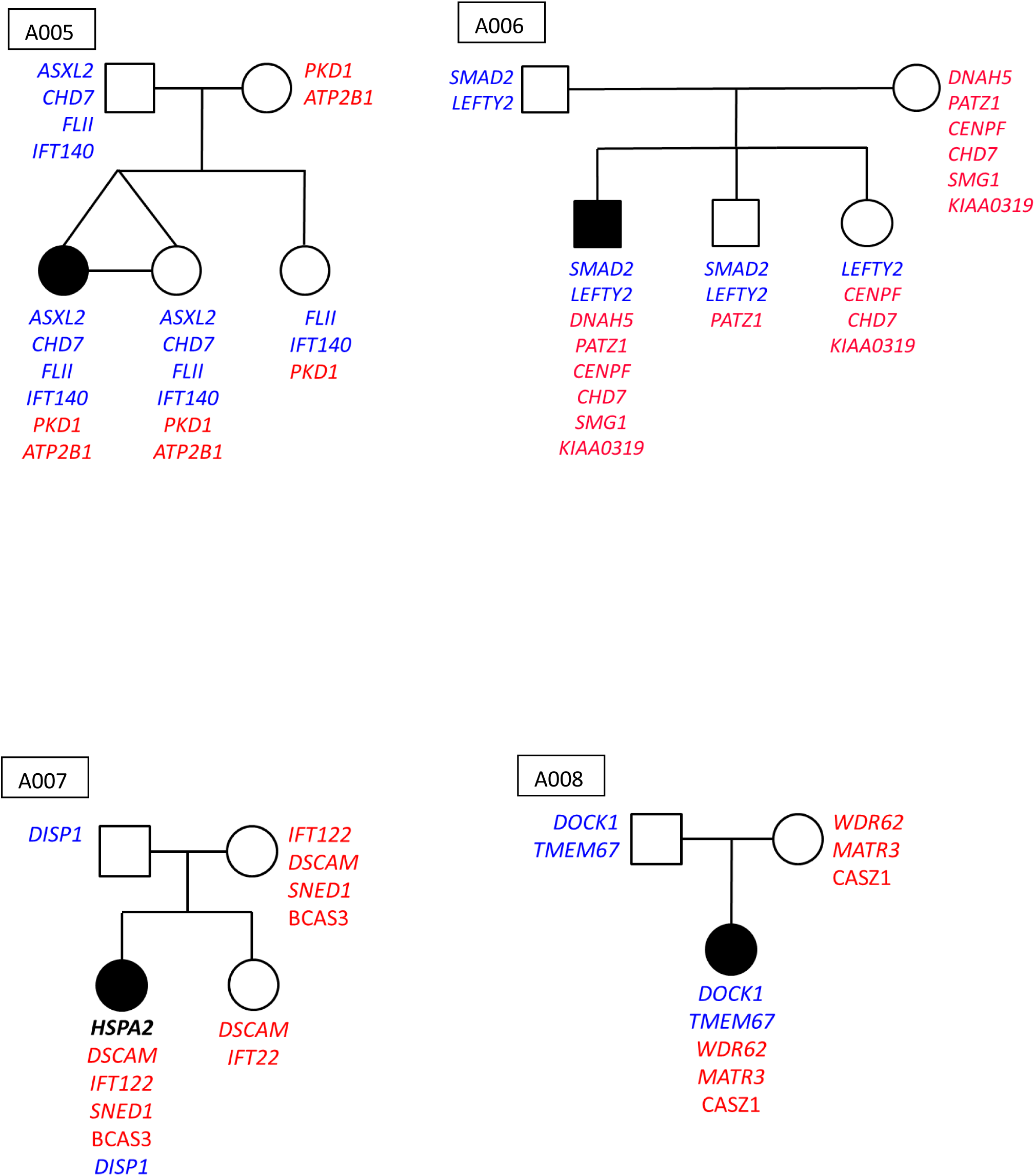
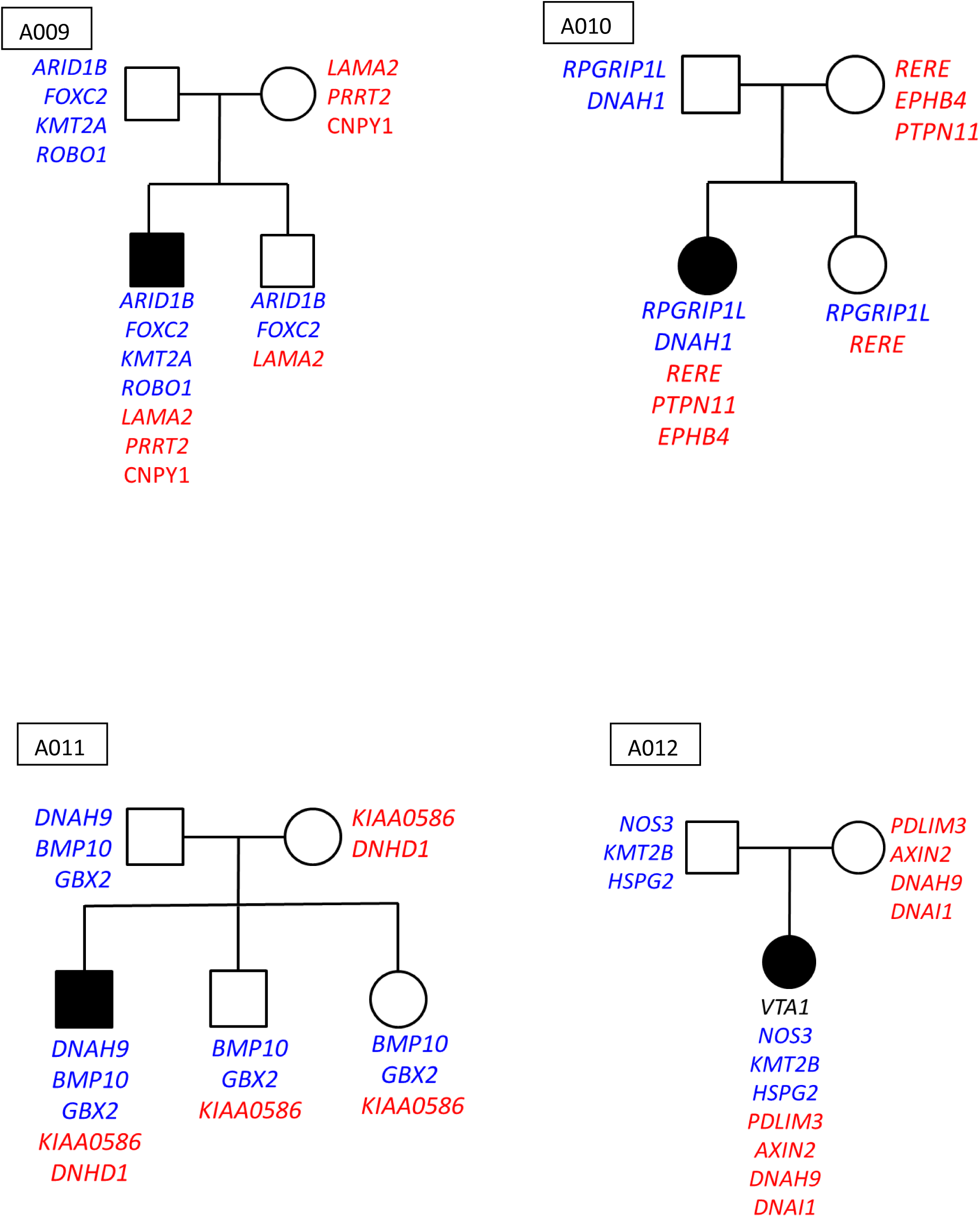
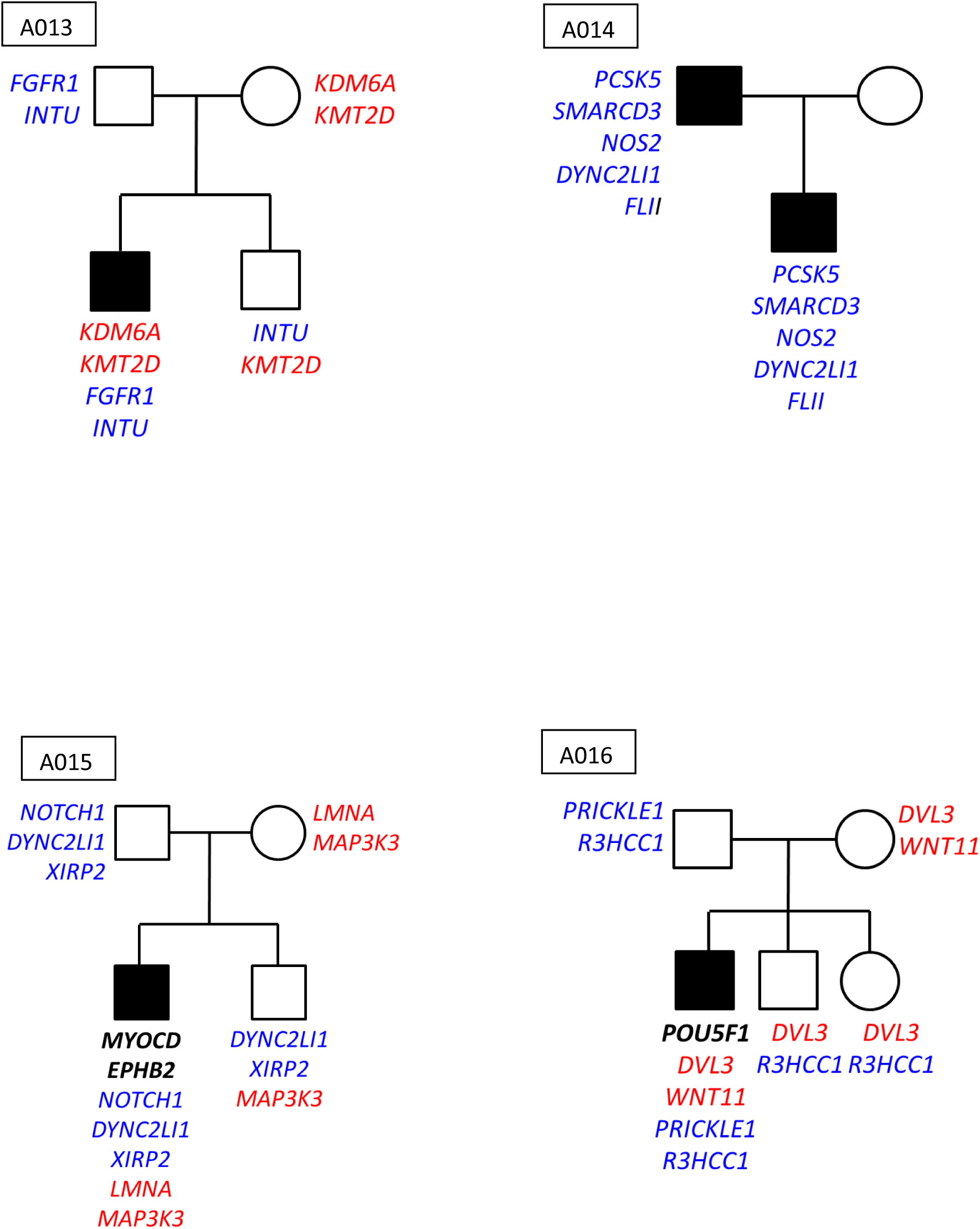
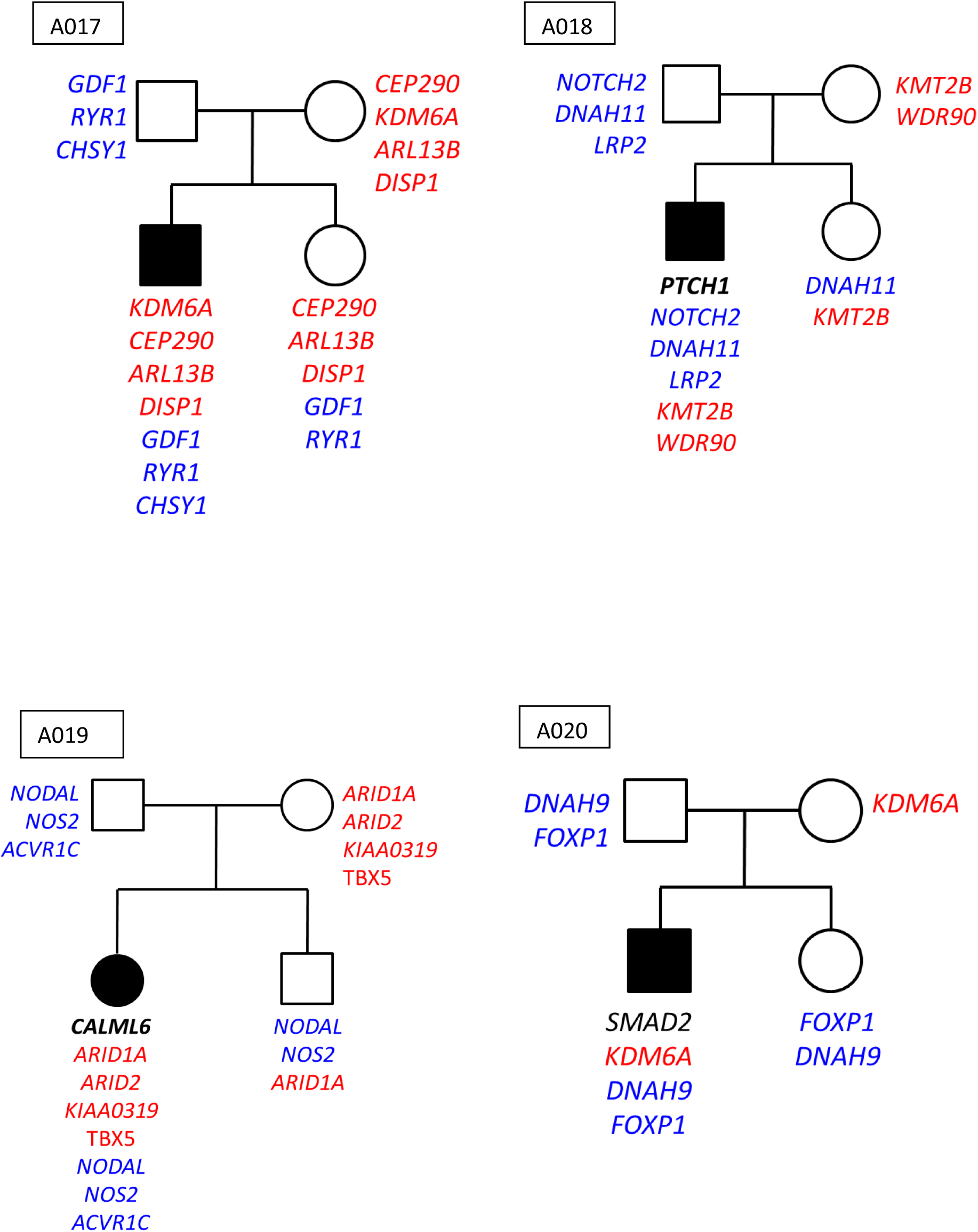
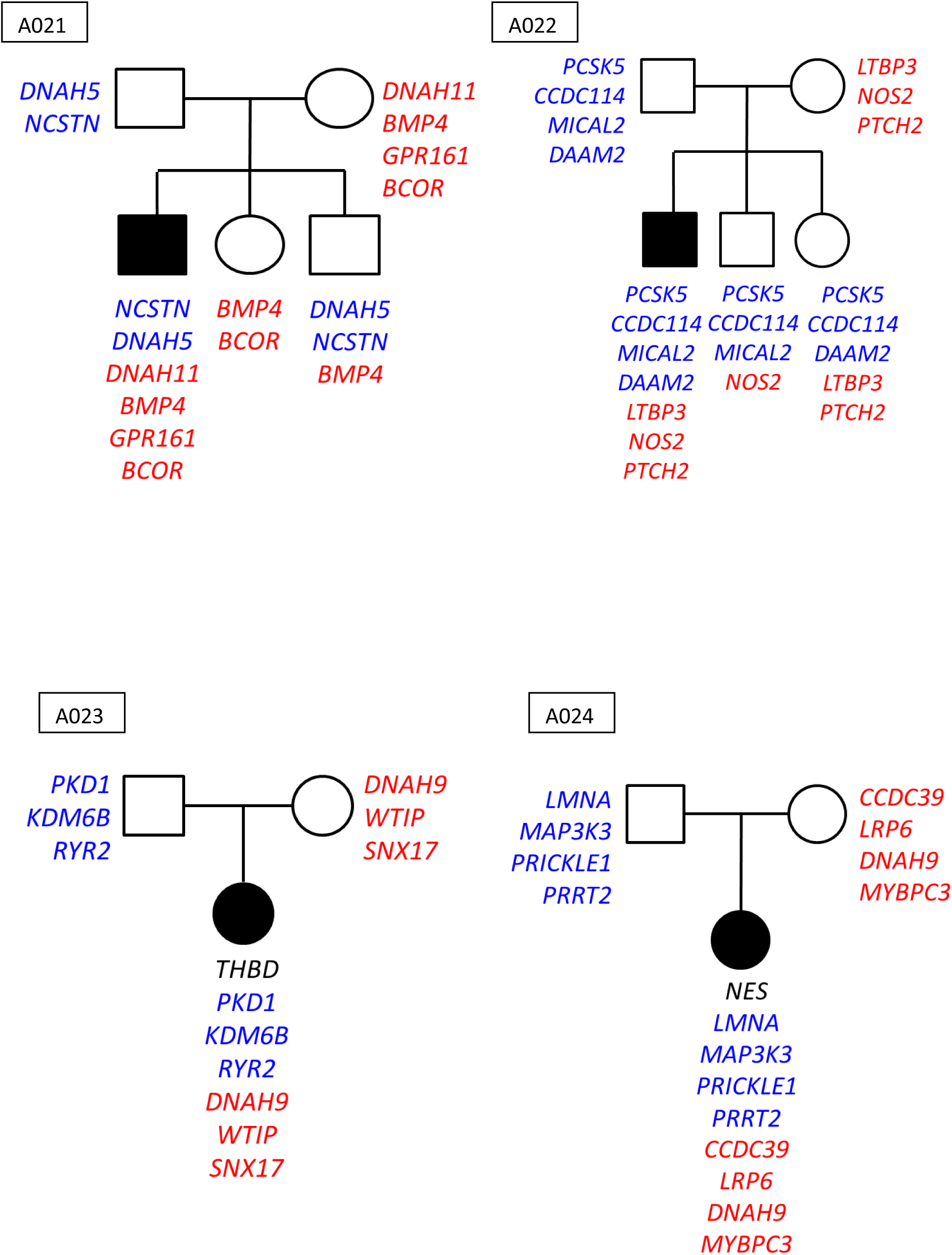
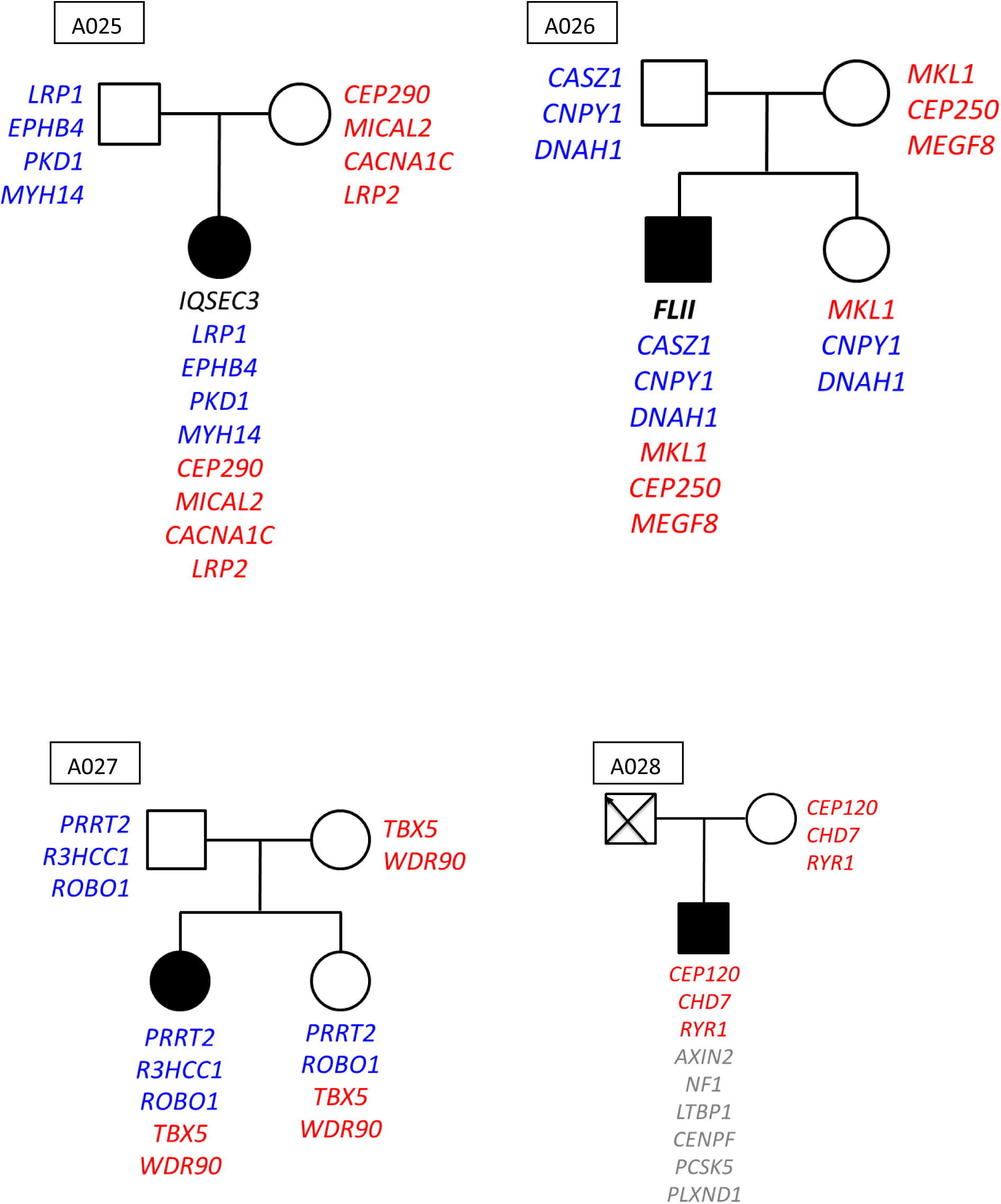
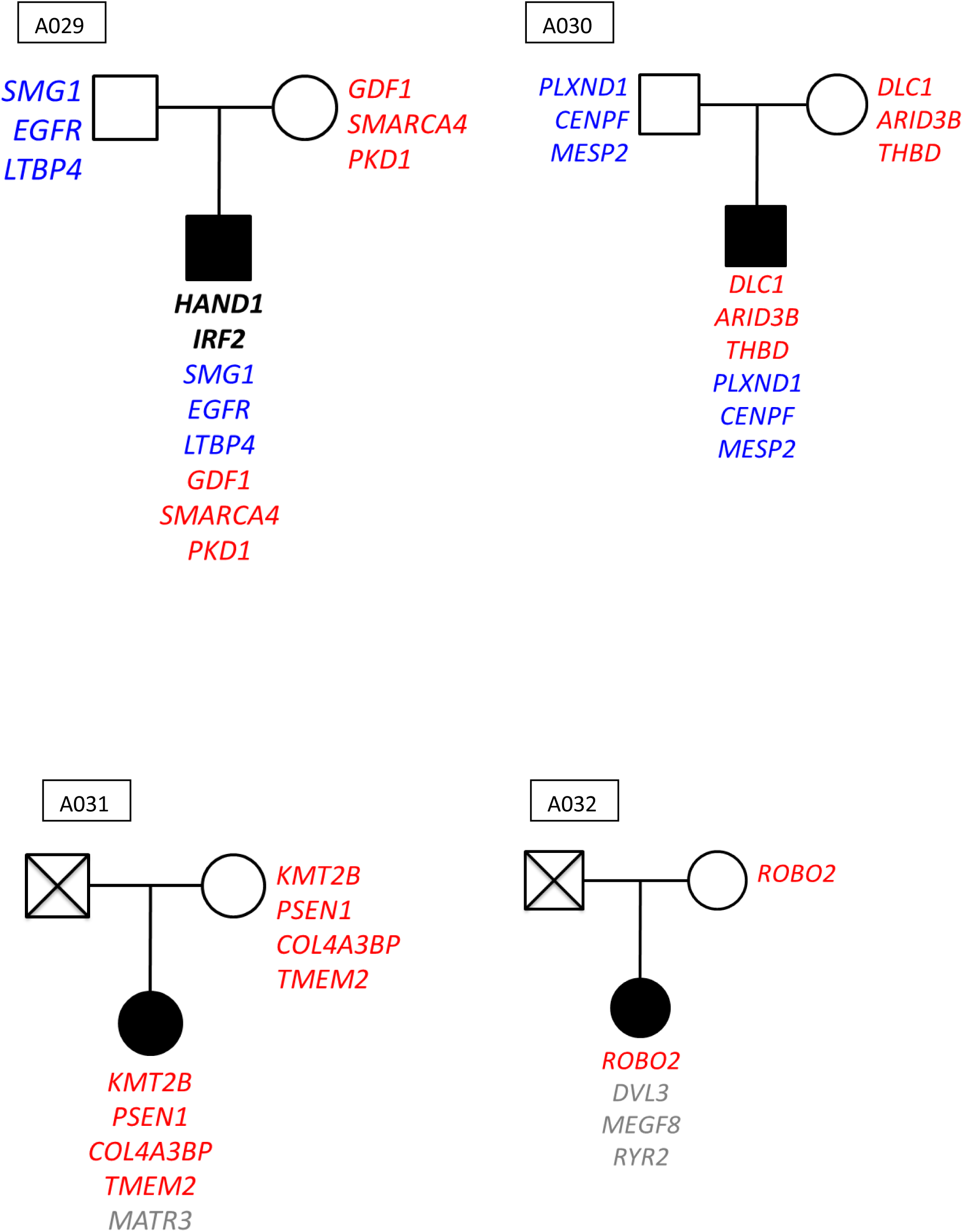
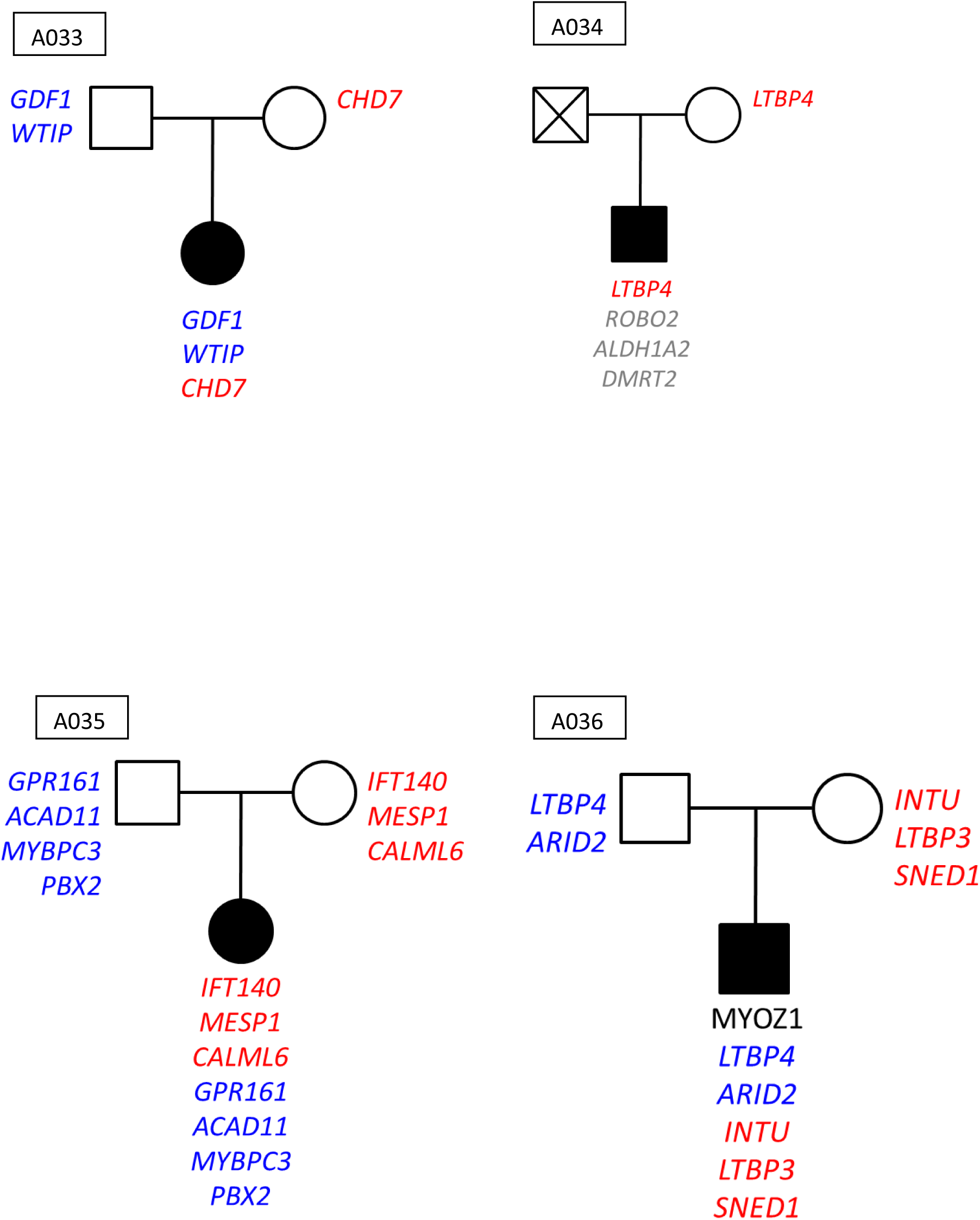
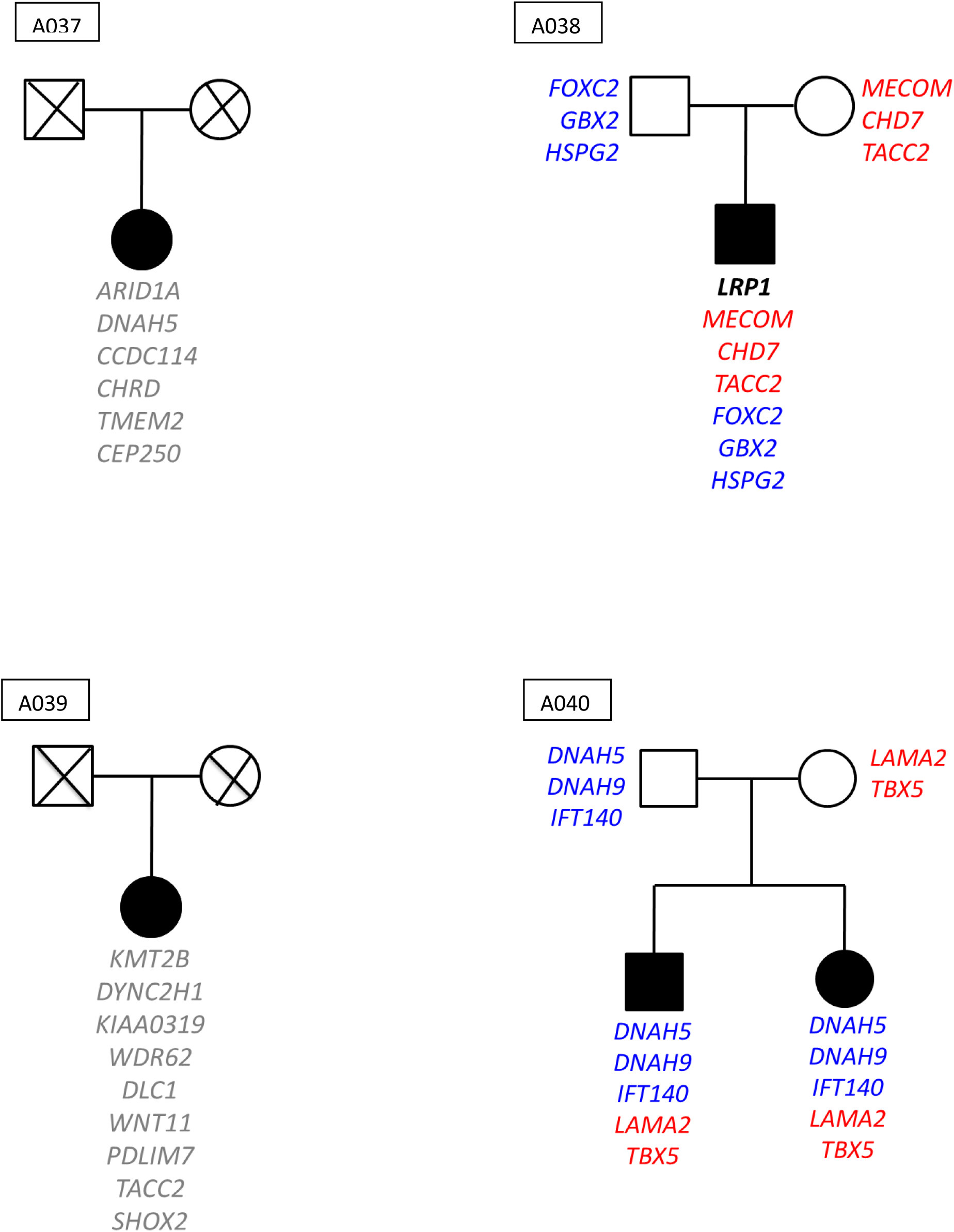
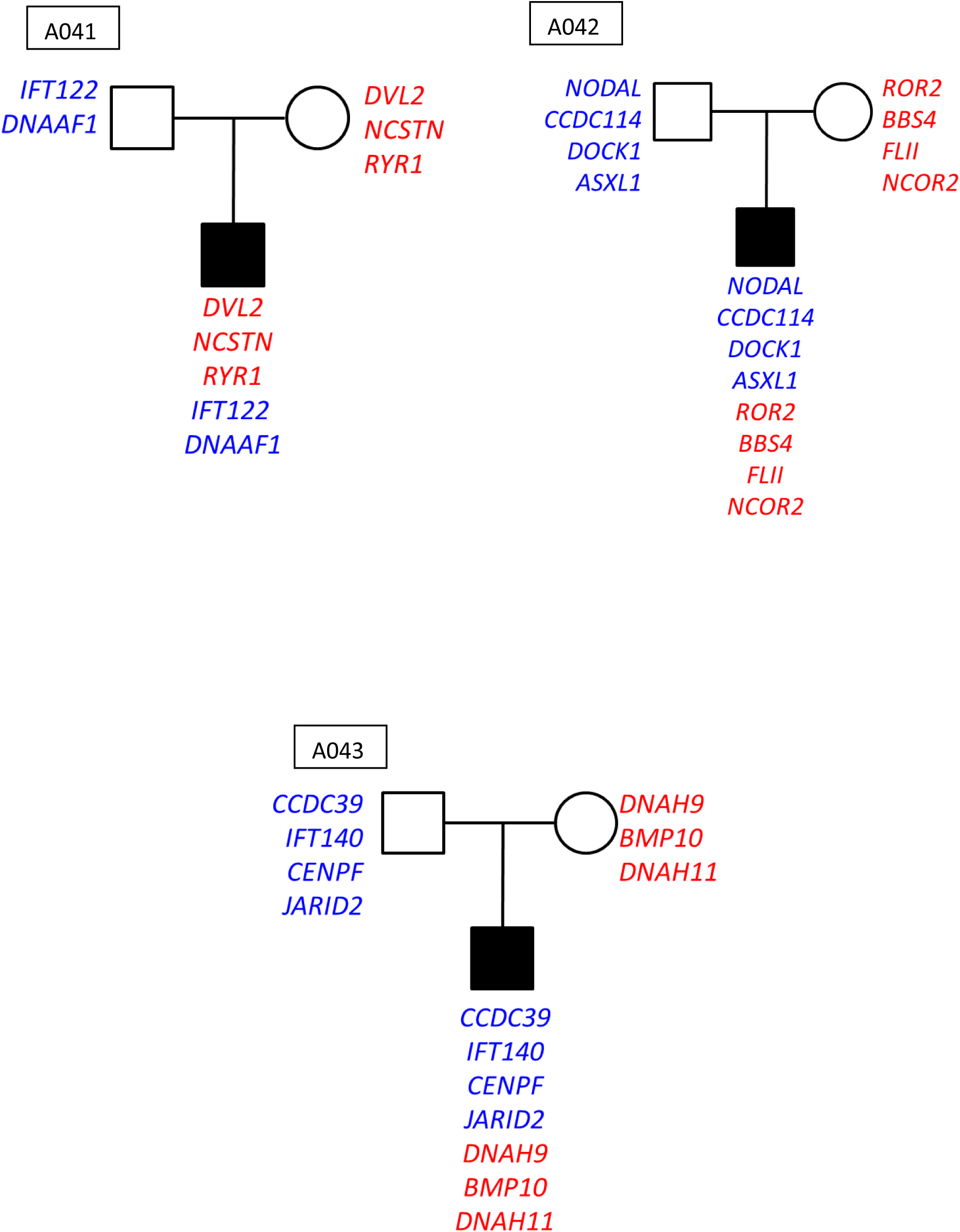
pedigrees of the 43 CCTGA families with the genotype of cases, parents and siblings. Color code: blue: variants inherited from the father. Red: variant inherited from the mother. Black: *de novo* variant. Grey: unknown inheritance

To support the hypothesis of a preferential transmission of variants in candidate genes from parents to sick children, we performed a family analysis with RV-GDT (Rare Variant Generalized Disequilibrium Test).

There was a significant transmission of rare variants in the 156 genes to the sick child (p=0.026).

To validate that these 156 genes are indeed linked to the disease, we tried to replicate our results on a German cohort of 86 index cases of non-syndromic CCTGA, and 5163 controls.

We used a logistic regression on the number of variants within the group of genes, according to the type of variants (PTV=protein truncating variant and PAV=protein altering variant).

We found a significant enrichment (*p value*=0.04, OR=1.5 for MAF 1%) in the replication cohort of PTV variants in the 156 genes of interest.

**Table (5)** summarizes these results.

**Table 5:** Replication cohort : statistical analysis.

| Cluster name | MAF | Type of variant | Total cases | Total controls | N_hets_cases | N_hets_controls | Nb var/cases | Nb var/controls | p_value | Odds radio |
| --- | --- | --- | --- | --- | --- | --- | --- | --- | --- | --- |
| **Geneset-156** | 1,00<sup>E</sup>-02 | PAV | 86 | 5163 | 479 | 31417 | 5,57 | 6,09 | 0,03 | 0,92 |
| **Geneset-156** | 1,00<sup>E</sup>-02 | PTV | 86 | 5163 | 18 | 716 | 0,21 | 0,14 | 0,04 | 1,5 |
| **Geneset-156** | 5,00<sup>E</sup>-03 | PAV | 86 | 5163 | 362 | 23776 | 4,21 | 4,61 | 0,05 | 0,92 |
| **Geneset-156** | 5,00<sup>E</sup>-03 | PTV | 86 | 5163 | 17 | 660 | 0,20 | 0,13 | 0,04 | 1,54 |

## Discussion

CHD is a large global problem in child health ^24^. Progress in the diagnosis and management of congenital heart disease is leading to reduced mortality and increased prevalence of CHD in adults. It is estimated that currently 85% of children with a complex CHD will reach adulthood. The population of adults with CHD is growing approximatively 5% / year ^25^. This highlights the need to lay a molecular foundation for these diseases. Basically, they represent the first step before physiopathological studies that will allow better understanding of the normal and pathological cardiac development leading to the main anomalies. Understanding the underlying genetic, molecular, and cellular mechanisms of CHDs will have real consequences for clinical practice, such as improving diagnosis, improving guidelines before surgery, and improving genetic counselling.

In this study, we sequenced a cohort of 44 cases of a complex and rare CHD, the congenitally corrected transposition of the great arteries, by whole genome and whole exome sequencing. Our data contradict a monogenic mode of inheritance, favouring a polygenic origin of CHD. In the majority of cases of our series, the parents harbour a set of susceptibility alleles for the disease that are inherited by the offspring in a combination that confers the risk of CCTGA.

Evidence exists for both inherited as Mendelian traits and *de novo* mutations associated to CHD. Yet, large multigenerational pedigrees demonstrating Mendelian inheritance are exceptional. Autosomal recessive inheritance of CHD has also been documented and these most extensively in populations with a high degree of consanguinity. Most of the reported multigenerational pedigrees of X-linked and dominantly inherited CHDs include either non-lethal CHDs such as atrial septal defect or those with variable phenotypes that include survivable CHDs and more severe manifestations of the same spectrum ^26^.

Up to now CHDs most often occurs sporadically, suggesting a role of *de novo* mutations in their pathogenesis. In 2013, the Paediatric Cardiac Genomics Consortium performed whole exome sequencing of 362 parent-offspring trios ^27^. This study found *de novo* point/insertion-deletion mutations in several hundred of genes that collectively contribute to at least 10% of severe CHDs. Another study reported by Bruencker et al in 2017 ^28^ and encompassing to date the largest genetic investigation of a single CHD cohort with 2871 probands, shows that the genetic contribution of *de novo* variants in CHDs was 8%. These elegant study do not demonstrate any causal relationship between the identifying variants and the occurrence of a CHD. Another hypothesis for this frequent sporadic occurrence of CHD is the limited role of genetic variations in causing the risk of CHD in the general population. This hypothesis is, with a very high probability, the reason for the scarce association of CHD with single gene mutation. A more holistic approach of the development of CHD should be developed without excluding the role of genes variations but considering that the cardiac morphogenesis would be disturbed when a combination of “cardiac” gene variations will exceed a certain threshold. This is the polygenic hypothesis for the cause of CHD. The hypothesis of polygenic inheritance of CHD is not new and is supported by the landscape of recurrence both in proportion of all CHDs’ incidence, and in the viewpoint of anatomical concordance. The number of genes’ variations in different “cardiac” genes may depend on the population considered. Further, it is also obvious that the set of genes to test to support this hypothesis is arbitrarily defined, and this is a limitation for describing the associations of genes conferring a risk for CCTGA. Here, we chose to have a minimal set of three “cardiac” genes to support the probability of an inherited combination ≤1%.

We thus built a set with 156 “cardiac” genes. Variants identified in these genes are characterized by their damaging effect and there shared role in heart development and/or have an embryological cardiac expression. Many interact with each other in the same signalling pathways. For most of them, mouse models show that mutations of these genes are associated with anomalies in the heart looping and the left-right asymmetry.

We managed to demonstrate that the CCTGA follows a complex polygenic model of inheritance, which consists of addition of susceptibility alleles inherited from the both healthy mother and father and that finally leads to a mutation burden in genes playing a role in heart morphogenesis in the affected sib.

We finally show through family segregation studies that a disease associated combination of variants is not replicated in healthy individuals in the index family.

Because of the complexity of our genetic polygenic model, no functional analysis could be conducted.

## Conclusion

Our data contradict a monogenic mode of inheritance, favouring an oligogenic origin of CCTGA, where both parents harbour genetic predisposition variants for the disease (susceptibility alleles) and both transmit this load of variants to the affected offspring, explaining the low recurrence risk.

## Data Availability

All data produced in the present study are available upon reasonable request to the authors

## Acknowledgements

We thank the families for their participation. We thank Anna PELET for technical assistance. This work was supported by grants from the Agence Nationale de la Recherche (ANR-Heart Asymmetry) and the Fondation pour la Recherche Médicale (FRM).

## Ethical and Legal Issues

I- For the human DNA studies, a particular agreement for the present study has been obtained with our Paris Descartes University (Île-de-France II) Ethical Committee in January 2010 (Comité de Protection des Personnes (CPP) agreement N° AOM 95224, P959892), according to which, each person involved in the study signed an informed consent; as for under-age individuals, the consent was signed by the parents as required by national laws. For obtaining written informed consent, each participating family has received an explanation (both oral and written) about the aims and objectives of the study.

II- Biobanking of patient materials (e.g. blood DNA) is achieved in our ISO9001-certified DNA1bank, within the Imagine building, in accordance with national, and international rules. The leukocyte DNA storage and automated extraction was performed at the Necker CRB-ADN bank, Centre de Resources Biologiques (CRB-ADN). DNA is stored in two aliquots, under anonymous codes. Of important note, our ongoing CARREG cohort sampling, has obtained an agreement from the CNIL (Commission Nationale de l’Informatique et des Libertés).

