## Supplemental figures for "Genetics of Congenitally Corrected Transposition of the Great Arteries: next generation sequencing shows a mutation load effect for 156 genes involved in cardiac patterning"

Supplementary figure 1: PCA of the primary cohort

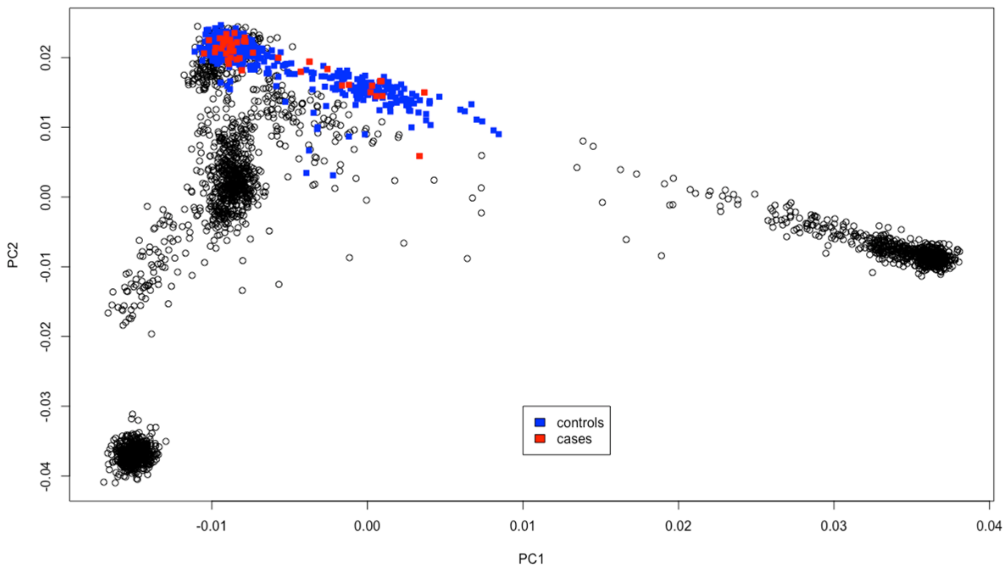

Supplementary figure 2: Comparison of synonymous variants in patients and controls

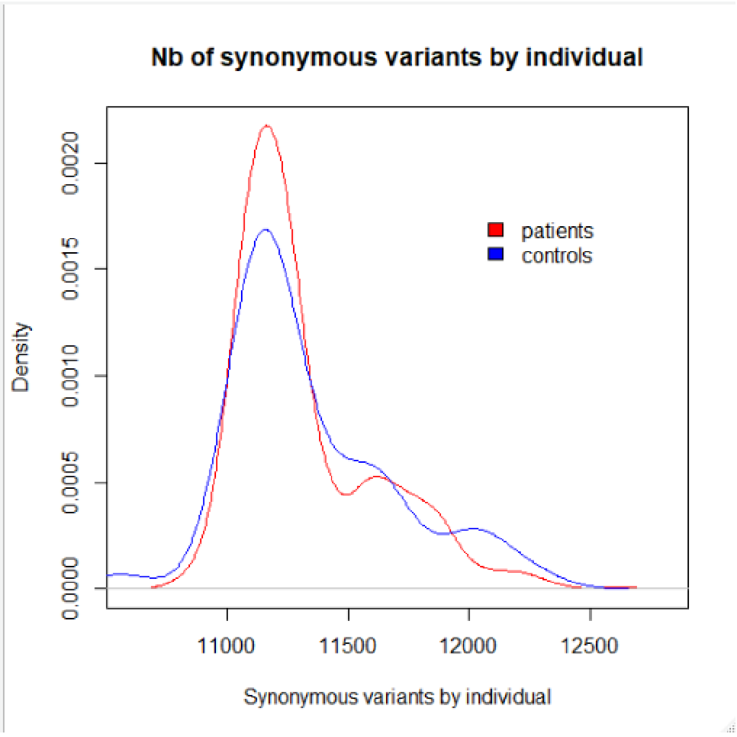
